# SARS-CoV-2 in 30 Months, Indonesia’s Data Tells: Study from a Reference Laboratory in North Jakarta and Its Reflection for Regional to National COVID-19 Situation

**DOI:** 10.1101/2023.07.27.23293237

**Authors:** Maria Mardalena Martini Kaisar, Tria Asri Widowati, Helen Kristin, Sheila Jonnatan, Sem Samuel Surja, Enty Tjoa, Venna, Jullyanny Waty Wijaya, Anita Devi K. Thantry, Ivonne Martin, Soegianto Ali

## Abstract

Atma Jaya Catholic University of Indonesia (AJCUI) COVID-19 Laboratory has become a reference testing site, which contributed to prevent the SARS-CoV-2 transmission in Indonesia. Through the LSSR and CARE, the Indonesian government has implemented moderate yet arguably successful policies to combat this pandemic. This study aims to assess and strengthen public health management while enhancing our understanding of the dynamics of COVID-19. We analyzed the correlation between policies enforced in controlling COVID-19 from July 2020–December 2022 to the positivity rate and viral intensity. AJCUI, DKI Jakarta, Indonesia demonstrated a similar trend in COVID-19 prevalence. Government policy on mobility restrictions has substantially reduced the positivity rate in Indonesia within the period of study. Our study interpreted that the Ct value in the positive case population of AJCUI data correlated positively with DKI Jakarta and Indonesia; subsequently, it has the potential to serve as an early warning for an anticipated wave. Despite the coverage of vaccines, AJCUI and DKI Jakarta positivity rates are shifting due to evolving virus variants. Altogether, the comprehensively recorded data would enable an understanding of COVID-19 dynamics, serve as a model for unprecedented disaster and public health management in general.

## 1. Introduction

At present, although the COVID-19 pandemic has been alleviated, its transmission continues across the entire globe. In December 2019, a novel coronavirus-caused infectious outbreak emerged in Wuhan, Hubei Province, China (2019-nCoV). In response to its rapid spread, on March 26, 2020, the World Health Organization (WHO) declared a COVID-19 pandemic. Accordingly, WHO published six ranked strategies for governments to implement regarding the epidemic: expand, train, and deploy healthcare workers; implement systems to find suspected patients; increase production and availability of tests; identify facilities that can be transformed into coronavirus health centers; develop plans to quarantine cases; and refocus government efforts on eradicating the virus were applied as strategies [1].

Reducing and delaying the epidemic’s peak is necessary. Uncontrolled measures will rapidly increase the number of cases, cause them to peak prematurely, and necessitate an expansion of the response capacity of the healthcare systems. In contrast, early implementation of stringent control measures will assist in reducing the number of cases, delaying the peak, and requiring significantly less capacity from healthcare systems. This concerning situation forces each country to ramp up strategies for combating this worldwide epidemic.

At the beginning of the COVID-19 pandemic, testing, tracing, and treatment were some of the preferred methods to curb the spread of the disease, including in Indonesia [2]. However, due to the widespread nature of the disease, tracing the contacts of the confirmed cases would not be an easy task to be implied. A different approach to an early warning in the health system should be in place, and we propose utilizing cycle threshold (Ct) values obtained from laboratories as an early warning. This kind of early warning allows us to take cautious measures, which could reduce the number of casualties, as we experienced during the emergence of the Delta variant of SARS-CoV-2.

Indonesia is the fourth-most populous nation in the world. In addition, Indonesia’s geographical situation consists of more than fifteen thousand islands [3].Consequently, it is expected to suffer significantly for a longer duration of the pandemic compared to the less populous nations. In Indonesia, the first two confirmed cases of COVID-19 infection were reported on March 2^nd^, 2020 [4]. However, as of April 2^nd^, 2020, marking one month since the first two cases were announced, there were already 1,790 confirmed cases, 113 new cases, 170 deaths, and 112 recoveries. Indonesia experienced the first, second, and third waves of COVID-19 over the course of nearly three years [5]. Through the Large-Scale Social Restrictions (LSSR) Policy *(in Bahasa Indonesia: Pembatasan Sosial Berskala Besar/PSBB)* and the Community Activities Restrictions Enforcement (CARE) Policy *(in Bahasa Indonesia: Pemberlakuan Pembatasan Kegiatan Masyarakat/PPKM)*, the government has implemented relatively moderate policies to combat this pandemic [6].

This study is the first comprehensive academic peer-review article based on data collected at the institutional, regional, and national levels where the policy responses are taken into account in reporting, analyzing, and evaluating current rapid responses to COVID-19 in Indonesia. Not limited to the current COVID-19 pandemic, policymakers are expected to be able to respond more effectively to future pandemics of a similar nature. Consequently, the purpose of this study is to assess and strengthen public health management by enhancing our understanding of the dynamics of the COVID-19 pandemic. In addition, we would analyze the correlation between policies enforced in controlling the spread of COVID-19 from July 2020 - December 2022 by looking at the fluctuation of the positivity rate and analyzing the correlation between the viral concentration of the positive cases and the positivity rate based on the data from our COVID-19 laboratory at Atma Jaya Catholic University of Indonesia, which may represent conditions in the wider DKI Jakarta area and in some cases, extend to Indonesia.

## 2. Methodology

As of June 20, 2022, the National Institute of Health Research and Development, Indonesian Ministry of Health (Litbangkes), had registered 936 COVID-19 Examination Laboratories in Indonesia, including 141 laboratories in the Special Capital District of Jakarta (in Bahasa Indonesia: Daerah Khusus Ibukota/ DKI), and 21 COVID-19 of the testing facilities are located in North Jakarta which is responsible to serve 146.66 Km^2^ area in Jakarta [7]. With the requirement of at least Bio-Safety Laboratory Level 2 (BSL-2) and a competent team, the Atma Jaya Catholic University of Indonesia (AJCUI) COVID-19 Laboratory, located in North Jakarta, began to operate on July 16^th^, 2020, with the Enhanced BSL2 standard. Thus, this laboratory became one of the first laboratories to join forces for diagnosing COVID-19. Every six months, the National Institute of Health Research and Development, Indonesian Ministry of Health, conducts an External Quality Monitoring Validation Test (in Bahasa Indonesia: Pemantapan Mutu Eksternal (PME)) to ensure each COVID-19 laboratory service is operating accurately and effectively thus the results from the laboratory can be certified valid.

### 2.1 Data Collection

Data extraction and collection were performed monthly from July 2020 to December 2022, obtaining 76,762 COVID-19 testing results from the AJCUI. We extracted the number of examined samples, positive samples, cycle threshold (Ct) values, genders, and ages from the AJCUI-received specimens. Atma Jaya COVID-19 Laboratory has recorded not only the qualitative results of administered specimens but since August 2020, has also documented the Ct value data of the positively tested specimens generated from Reverse Transcription-quantitative Polymerase Chain Reaction (RT-qPCR). Thus, the recorded Ct values can be used for further analysis. In addition to the data from the AJCUI, we also extracted data on the number of daily tested and positive samples from DKI Jakarta and Indonesia, as available on the official websites. There were 12,590,998 data from DKI Jakarta and 106,648,851 data on Indonesia’s COVID-19 cases obtained from DKI Jakarta government-specific website for COVID-19, Jakarta Tanggap COVID-19 (https://corona.jakarta.go.id/id) and Indonesia COVID-19 dashboard (https://infeksiemerging.kemkes.go.id/dashboard/covid-19), WHO COVID-19 Dashboard for Indonesia (https://covid19.who.int/region/searo/country/id), respectively. Furthermore, policy details are provided in **Supplementary File 1** obtained from the Cabinet Secretariat of the Republic of Indonesia’s website according to the executed decree (https://setkab.go.id/). Additional vaccination data of DKI Jakarta were obtained from the Ministry of Health Vaccination website (https://vaksin.kemkes.go.id/#/vaccines). Data on SARS-CoV-2 variants in Indonesia were retrieved from GISAID, Submission Tracker Global [8]. Collected data were extracted, stored in Microsoft Excel, analyzed, and visualized using GraphPad Prism 9.1.1 (GraphPad Software, La Jolla, CA, USA).

### 2.2 Data Analysis

Data analysis was carried out by comparing the COVID-19 positivity rate of retrospective specimens received by AJCUI with the data from DKI Jakarta and Indonesia, from July 2020 to December 2022. We divided the age data from AJCUI into seven groups: 0-10 years, 11-20 years, 21-30 years, 31-40 years, 41-50 years, 51-60 years, and more than 60 years old. We observed the trend of age groups as well as gender categories getting tested for COVID-19 with heatmap visualization. Simple linear regression was performed on the Ct value analysis in AJCUI to see whether the data is related to the fluctuation of COVID-19 cases in both AJCUI and DKI Jakarta. We calculated and analyzed the slope of regression (β), the 95% confidence intervals (95% CI), and the R squared (R²). For visualization, data was made into explanatory graphs. Within the period of data extraction, AJCUI COVID-19 laboratory detects SARS-CoV-2 based on RT-qPCR with differing detection kits that detect at least three to four targeted genes, including the housekeeping gene for the analysis control. The genes detected were ORF1ab, N, E, and RdRp. Positive samples with Ct value data were grouped into two, i.e., less than and 30 or more. The readout would be below 30 if at least two genes showed values below 30 and vice versa.

## 3. Results and Discussion

### 3.1 AJCUI-DKI Jakarta-Indonesia data reveals a similar trend of COVID-19 prevalence

Monthly percentages of COVID-19-positive cases based on AJCUI data from July 2020 to October 2022 were higher compared to both DKI Jakarta and Indonesia’s data, as can be appreciated from **Figure 1** and **Supplementary File 2**. Those could be explained by some factors. First, at the time, the testing at commercial laboratories was individually paid with relatively high costs, which not everyone could afford, whereas DKI Jakarta’s data and Indonesia’s would include not only commercial laboratories but government health care diagnostic laboratories that would offer free tests despite delays in the release of results [9]. Second, uneven testing coverage, particularly across Indonesia, due to the availability of COVID-19 diagnostics centers by means of a proper laboratory for handling airborne viruses such as SARS-CoV-2, which require a minimum BSL-2 facility along with limited reagents to perform the test [10]. These first and second factors were the primary concerns in the early period of COVID-19. The testing ratio of major cities, such as Jakarta, will differ from that of other cities, such as those on Java Island. The third factor is affected by people’s self-awareness. Individuals who came directly to the laboratory for testing were quite aware and expected to have a higher probability of being positive due to their symptoms or past exposure to confirmed cases. Despite the difference in positivity rate across these three data sets, the positivity rate follows a similar trend in terms of its dynamics.

**Figure 1.**
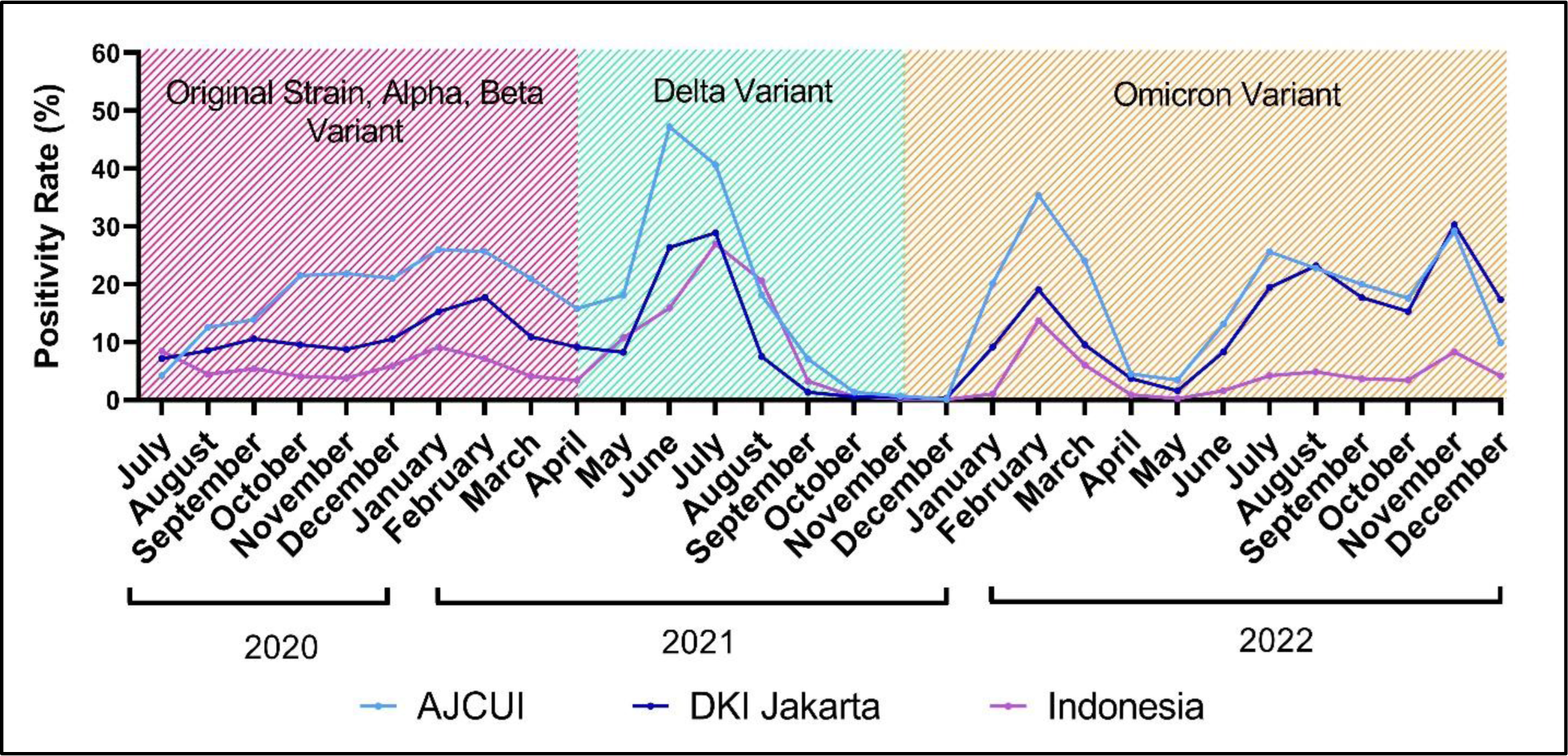
The COVID-19 dynamics prevalence from July 2020 to December 2022. The trend of COVID-19 displays for 30 months based on the prevalence data from AJCUI, DKI Jakarta (source: Jakarta Tanggap COVID-19 (https://corona.jakarta.go.id/id)), and Indonesia (source: Indonesia COVID-19 dashboard, (https://infeksiemerging.kemkes.go.id/dashboard/covid-19)) The shaded areas within the graph represent an approximation of the dominant SARS-CoV-2 variants. The colors red, green and orange correspond to the origin strain-Alpha-Beta, Delta, and Omicron variants, respectively.

A few occasions of a sudden increase in COVID-19 cases were observed in laboratories. In this instance, from **Figure 1**, we observed data from the AJCUI, which might represent cases from DKI Jakarta and Indonesia, as it demonstrates a comparable trend. As the majority of other countries experienced a surge of COVID-19-positive cases caused by so-called responsible variants [11]. The data from July 2020 to December 2022 revealed that there were three waves dominated by different SARS-CoV-2 variants with respect to the monthly period in Indonesia. **Figure 1** shows the first wave, represented by the red area, was nationally dominated by Alpha and Beta variants. The second wave was dominated by the Delta variant, depicted by the green area [12]. During this second wave, the positivity rate reached its peak in June 2021 for AJCUI and in July 2021 for DKI Jakarta and Indonesia. The Delta variant has been shown to have a higher transmission rate than the previous variants, even in fully vaccinated people, and it is estimated that the Delta variant is twice as contagious compared to the previous variants [13–15]. Originating from India, whose first case was found in October 2020, this strain transmitted with high growth rates in various countries, such as the United Kingdom, Hong Kong, China, Singapore, Russia, the United States, and Australia [16]. The third wave, denoted by the orange area, was dominated by the Omicron variant, resulting in two peaks of cases during the B4 and B5 subvariants in February 2021 and the XBB subvariant [17]. The peak of cases occurred in July, August, and November 2022 for AJCUI and DKI Jakarta, but nationally for Indonesia, the peak was prominent only in November 2022. From July 2020 to May 2022, the positivity rate for DKI Jakarta and Indonesia were comparable. However, from June to December 2022, DKI Jakarta data tend to resemble AJCUI data. Since AJCUI has not been in operation until July 2020, we cannot observe the case trend before that date.

The testing and positivity rate dynamics across all three datasets generally exhibited a similar trend in July 2021. Forty out of one hundred individuals tested positive for COVID-19 at AJCUI. Meanwhile, in DKI Jakarta and Indonesia, 29 out of 100 and 27 out of 100 of those tested were positive for COVID-19, respectively. However, between June and November of 2022, AJCUI and DKI Jakarta had a significantly higher positivity rate compared to the national data (**Supplementary File 1**). This was due to an increase in the number of positive cases as the testing rate decreased. In addition, between December 2021 and April 2022, there was an abrupt increase in the frequency of tests. This phenomenon was the consequence of travel regulations that required negative PCR testing, particularly for air travel.

### 3.2 Contrasting AJCUI testing and positivity rate towards parallel data from DKI Jakarta and Indonesia

During the initial stages of the COVID-19 outbreak, the cost of a PCR test was still prohibitively expensive, reaching IDR 2.5 million per test. In considering the substantial cost of molecular testing for the general public, the Ministry of Health set a price limit of IDR 900 thousand. Due to the development of molecular test kits and the expansion of testing facilities, the price of this PCR has fallen significantly [18]. As part of the government’s effort to increase the testing rate for contact tracing, the cost of COVID-19 testing utilizing RT-PCR was lowered by approximately threefold on October 27, 2021. We assumed that this might account for the increase in specimens tested in DKI Jakarta and Indonesia laboratories after October 2021 [19].

The third wave, which occurred during the domination of the Omicron variant, was indicated by a spike in examinations and positive cases, which can be observed from December 2021 to March 2022. In Indonesia, the first case of Omicron was reported in early December 2021. Since then, the Omicron variant has infected a growing number of people [20]. Yet, starting in December 2021, many individuals began to travel locally and internationally for vacation at the end and beginning of the year. Prior to their trip and following their arrival, they are subsequently tested for COVID-19. Many individuals seek medical attention whenever they exhibit symptoms of COVID-19, presumably out of concern that another wave with similarly severe symptoms will emerge, as was the case with Delta [21].

In Indonesia, starting from late July 2022, those vaccinated up to the third dose (booster) are no longer required to present negative COVID-19 test results to travel by land or air within the country. However, individuals who have only received the second dose of the vaccine must provide a negative antigen or RT-PCR results for COVID-19. Despite new omicron sub-variants continuing to be discovered, the availability of the more affordable antigen and other COVID-19 tests, increasing herd immunity due to higher vaccination coverage, and a more relaxed travel regulatory policies contribute to a decline and flattening of the testing rate, which may also contribute to the identification of positive cases.

### 3.3 Testing and positive cases dominated by the high mobility age groups

The age data were obtained solely from the AJCUI COVID-19 Laboratory, which majority was a paid service provided by the Atma Jaya Hospital. Samples were retrieved from the emergency department, inpatients, and drive-thru swab services. At the drive-thru service, the specimens were received from patients with travel or tracing purposes. Accordingly, from **Figure 3a**, we can conclude that people of younger ages (the twenties to fifties) with higher mobility dominated the age pool. The percentages for each age group were as follows: 21–30 (29.27%); 31–40 (22.30%); 41–50 (16.55%); 21–50 (68.11%). The higher proportion of particular age groups being tested was in line with the positivity prevalence being higher in those age groups (**Figure 3b, Supplementary File 3**). This trend occurred consistently for the data series collected from July 2020 to November 2022.

**Figure 2.**
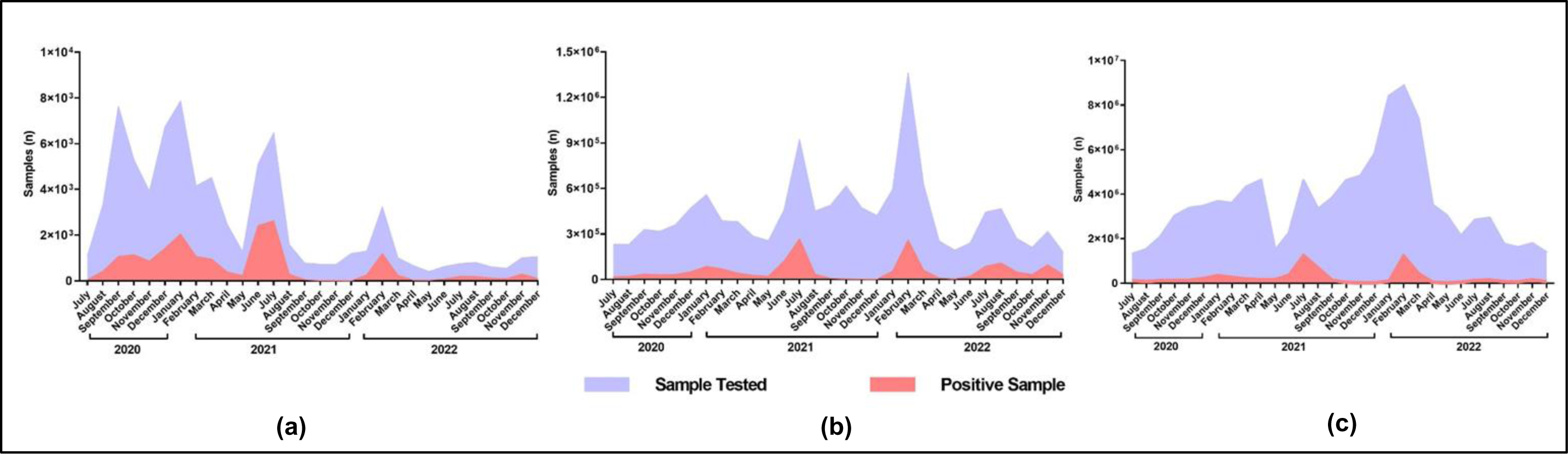
The trend of testing and positivity rate of COVID-19. The graphs depict the data recorded from July 2020 to December 2022. The purple shaded areas represent the number of tested specimens, overlayed by the red areas which represent the number of positive specimens detected by RT-qPCR. The trend of both testing and positivity rate showed for (a) AJCUI, (b) DKI Jakarta (source: Jakarta Tanggap COVID-19 (https://corona.jakarta.go.id/id)), and (c) Indonesia (source: Indonesian Ministry of Health social media page).

**Figure 3.**
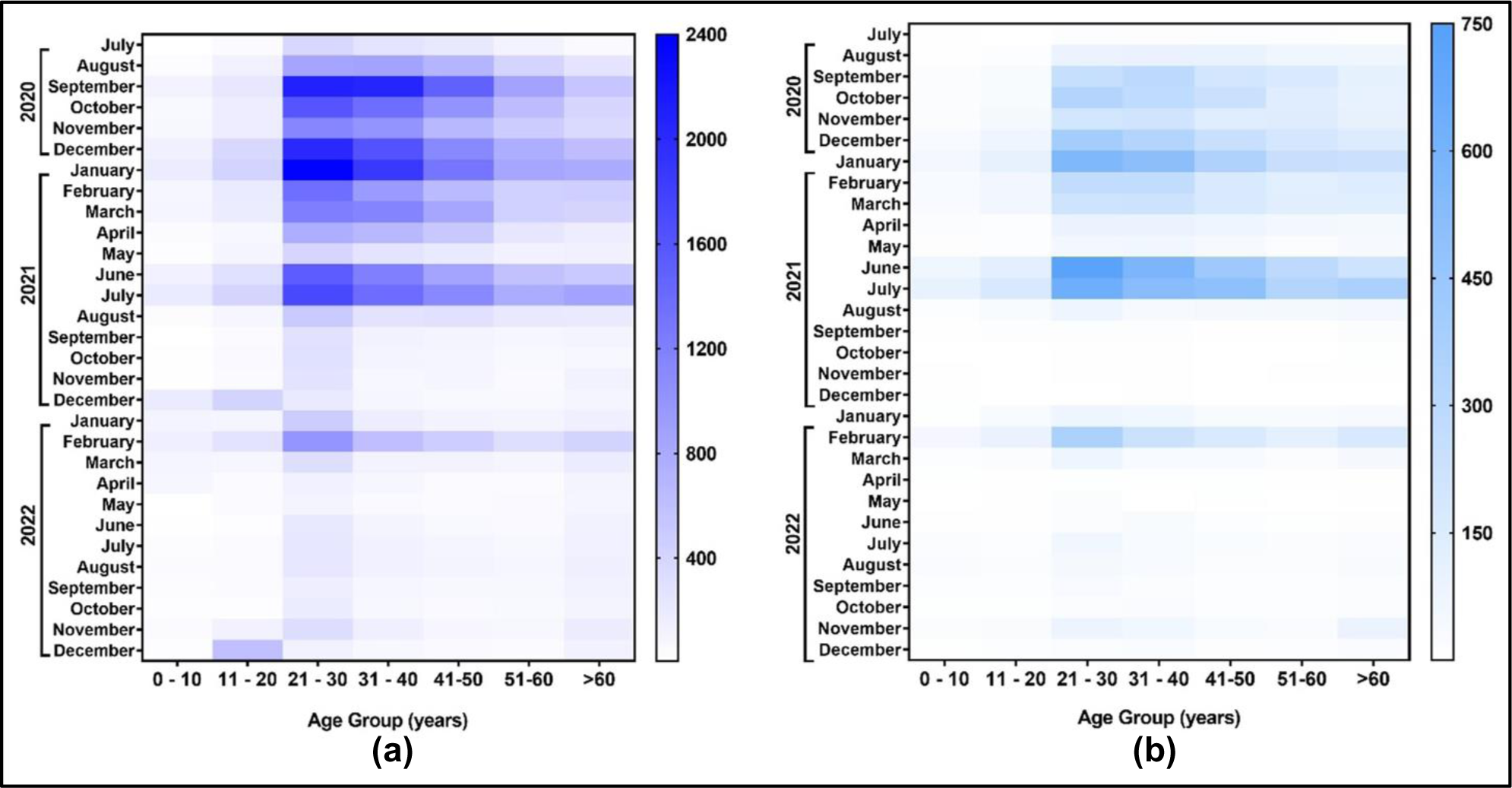
The heat maps of age groups stratifications to COVID-19 testing and positivity administered to AJCUI. The data shown was based on specimens collected from July 2020 to December 2022. The darker the color, the higher the represented number. The age group stratification heat maps compare (a) the testing rate to (b) COVID-19 confirmed cases by RT-qPCR.

Adults are typically more conscious of being swabbed for security purposes than children. Consequently, it was inevitable that the older group would dominate the age pool. Similar data were presented during the first wave of the COVID-19 pandemic in Italy; older adults, excluding those with a clear-cut clinical indication, were less likely to be tested for SARS-CoV-2 infection compared to younger individuals [22]. A study by researchers at Touro University and its affiliated New York Medical College discovered that the rate of infection was significantly higher among younger individuals compared to older adults, which might be due to the lack of social behavior in the COVID-19 protocol [23]. In addition, during the surge of COVID-19, several companies in Jakarta required their employees, which are in the productive age range of 21–50 years old, to submit negative PCR swab results prior to work from the office. Therefore, younger individuals frequently undergo COVID-19 testing [24,25]. From the beginning of the COVID-19 pandemic, the Indonesian government has declared some essential sectors, such as healthcare-related services (hospitals, clinics, pharmacies, and medical supplies), manufacturers (personal care, hygiene, and medication), grocery and convenience stores, financial and banking sectors, energy-related industries, as well as security (police and military forces), to continuously operate, thus supporting the public’s needs during the pandemic. Similar policies, although to varying degrees, were implemented in other nations [26–28]. to ensure human survival. As stated previously, the productive age groups of the 20s to 50s comprised the majority of the sector’s workforce. Considering their mobility, it is reasonable to assume that these age groups were naturally subjected to the highest testing.

The percentage of positive cases in the AJCUI COVID-19 laboratory was categorized according to the age groups as follows: 1–10 (3.17%); 11–20 (5.63%); 21–30 (26.51%); 31–40 (22.80%); 41–50 (16.91%); 51–60 (12.42%); > 60 (12.49%) as shown in **Figure 3b** and **Supplementary File 3**. The age group between 21 and 30 has the greatest positivity rate. This data could be explained considering that the percentage of swab samples received by the hospital was dominated by this age group. The assumption that young people were testing themselves at the COVID-19 test site due to contact with positive individuals is also likely to be proven. From these findings, we can also conclude that higher mobility increases the risk of virus exposure. A previous study conducted in Florida has also suggested that individuals between the ages of 15 and 24 have a higher risk of contracting COVID-19 than other older age groups. In Minnesota, the incidence of positive cases is higher among those aged 10 to 24 than among those aged 65 and older [23].

The percentage of men and women that underwent self-testing at the AJCUI laboratory in total (positive and negative cases) was 50.3% and 49.7%, respectively, which is comparable as indicated in **Supplementary File 4**. Out of 17,840 positive samples, men account for 50.58% and women for 49.42%. The same holds true with the data on age correlation with both testing and positivity rate; this gender data is exclusive to the AJCUI Laboratory and cannot be a representation of DKI Jakarta or Indonesia. The reason is that both age and gender data were not available from the linked public sources that we used. Men are more likely to get infected and/or check themselves more regularly than women since they have frequent contact with infected individuals due to their higher mobility [29,30]. However, there are a number of reasons that may contribute to this disparity. Moreover, according to several studies, women showed more responsible attitudes concerning the COVID-19 pandemic than men. The careless attitude of men adversely affects their compliance with preventative measures such as frequent handwashing, face mask use, and stay-at-home directives [31].

### 3.4 Policies enforced effectively reduced the positivity rate

According to WHO standards, the severity of a pandemic situation is determined by the level of transmission and the capacity to respond to that level of transmission. The LSSR is a restriction regulation whose particulars are governed by each region and approved by the Minister of Health. Schools, shops, offices, shopping malls, and tourist attractions must be temporarily closed during the LSSR or until the number of COVID-19 cases can be managed in the available health facilities. However, several regions, including DKI Jakarta, excluded eight basic, necessary-related business sectors. The eight sectors are permitted to remain operational despite the implementation of the LSSR by the Provincial Government of DKI Jakarta. Health-related care and businesses, such as hospitals, clinics, and personal care manufacturers, are the first permitted to operate (i.e., disinfectants, soaps, etc.). Similarly, businesses in the food and beverage, energy (water, gas, electricity, and gas stations), and telecommunications industries were permitted to operate. In addition, financial and banking sectors, such as stock exchange, logistics activities, and product distribution, were permitted to operate [32].

The government of Indonesia formulated its policies based on a number of factors, including the number of positive cases, the death rate, the healthcare capacity in each region, and the weekly evaluation of the level of restriction. Following LSSR, the government introduced CARE, a new regulation that is quite distinct from LSSR. The central government directly establishes CARE limitation regulations for each region based on regional conditions. Level 4 CARE was applied when the transmission was dispersed, and the response capacity was insufficient. Level 3 was implemented when a community outreach situation had limited response capacity. If there was an imminent risk of inadequate health services, level 2 was executed when the situation had a low community incidence. As for level 1, it was applied when the transmission was very low and the capacity of healthcare was ample. Nevertheless, there were limitations in implementing efforts to prevent transmission, or transmission occurred but could still be controlled; level 0, which has never occurred in DKI Jakarta, is a situation in which there is no local transmission [33]. **Supplementary File 1** provides detail on implemented policies since the beginning of the pandemic to date, along with the release and implementation dates of the policies.

The success of LSSR and CARE was measured by the implementation of the health protocol, in addition to taking these policy-changing factors into account. Based on a national survey conducted by Saiful Mujani Research & Consulting (SMRC) in 2020, the citizens of Jakarta were able to reduce the transmission of SARS-CoV-2 by enforcing strict LSSR restrictions. Consequently, it can be assumed that LSSR has effectively contained the pandemic. Although LSSR was well implemented in Jakarta, the implementation was still lower in other provinces, such as East Java and South Sulawesi [34]. To address this situation and in an effort to create a coordinated policy controlled by the central government and not by local governments, the CARE policy was created. In addition, the policy shift is intended to loosen restrictions, allowing citizens to maintain their economic standing despite the LSSR’s strict mobility restrictions. Based on the declining death rate, CARE was successfully implemented in Jakarta, which became a model city for CARE implementation [35].

A case study conducted among university students in three Indonesian cities in January 2020, April 2021, and July 2021 revealed that the students’ health protocols and practices varied in frequency. A face mask covering the chin and nose was the most common health protocol among students while limiting mobilization and maintaining distance was the most violated [36].

When there was a significant increase in the positivity rate, the government implemented stricter policies, and when the positivity rate decreased, the government loosened these policies. The policy shift was also gradual, but its effect on the positivity rate was immediate. **Figure 4** highlights the DKI Jakarta policies in effect. During the early period of SARS-CoV-2 transmission, LSSR was implemented by the public in Indonesia due to the unpredictability of the viral spread [37]. However, a shift in regulation was initiated from LSSR to micro-scale activity restriction followed by emergency care considering the high number of COVID-19 cases [38,39]. The increase and the high number of COVID-19-positive patients may have been driven by the Delta variant, which has a higher transmission rate than prior variants and is considered to be twice as contagious as earlier variants [13–15]. Subsequently, on July 26, 2021, as a result of strict emergency CARE regulations, the number of confirmed COVID-19 cases declined, allowing the public to adjust their mobility according to CARE Levels 4 and 3 [40,41]. A month later, the implementation of CARE was loosened, signifying the global recovery and marking two years of the pandemic. Along with the stated vaccination program, the two least stringent restrictions, CARE level 2 and 1 policies, were implemented based on the percentage of COVID-19 cases [41]. Whenever a situation represents an increase in positive cases, Level 2 CARE is regulated, which is contradictory when positive cases indicate a decline, in which Level 1 CARE is operated.

**Figure 4.**
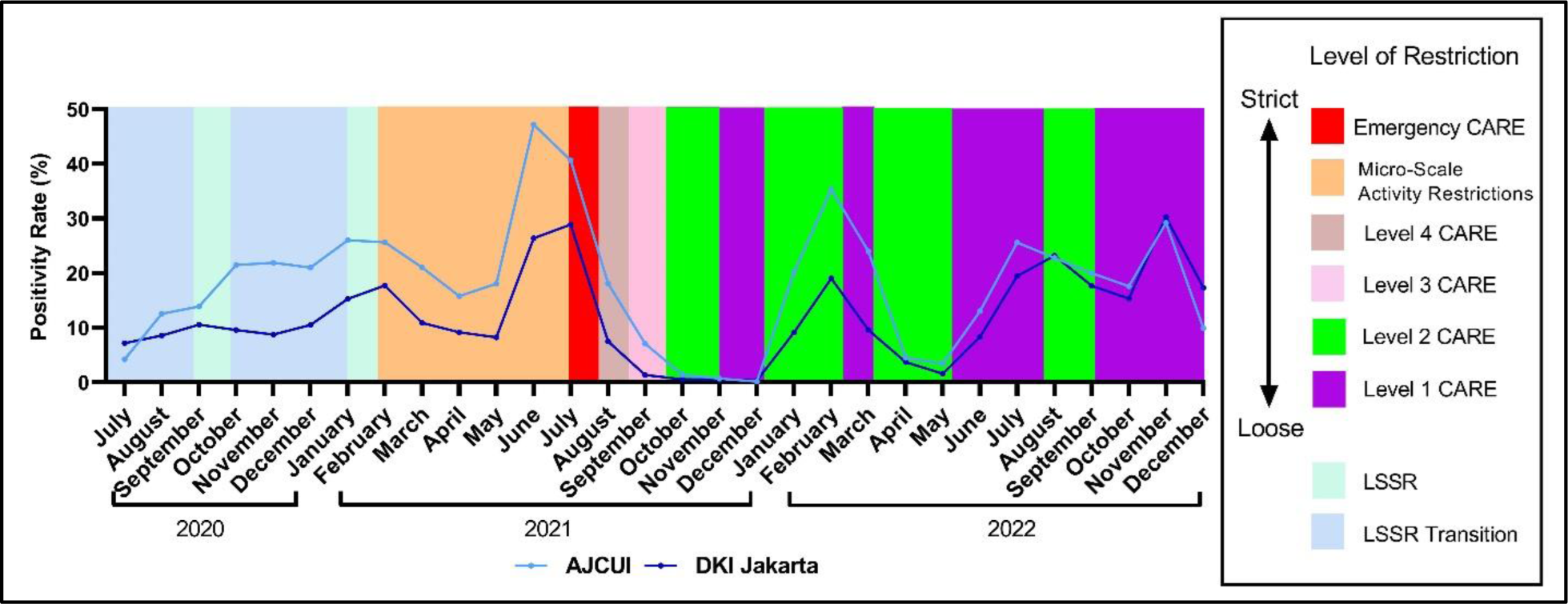
The periodic adjustment of policies enforcement according to the COVID-19 positivity rate. The two overlay lines represent the fluctuation of COVID-19 confirmed cases detected by RT-qPCR at AJCUI COVID-19 Laboratory (light blue line) and the data from DKI Jakarta (dark blue line) extracted for the period of July 2020 to December 2022. LSSR: Large-Scale Social Restrictions; CARE: Community Activities Restriction Enforcement.

### 3.5 Higher viral load corresponds to a greater proportion of positive cases and vice versa

Based on the rendered data from three sources, AJCUI, regional (DKI Jakarta), and national (Indonesia), comparable trends of positivity percentage and testing-to-positive-case ratio were observed. We further evaluated the correlation between the COVID-19 positivity rates at AJCUI and DKI Jakarta. As shown in **Figure 5a**, there was a positive correlation between the cases observed in AJCUI and those in DKI Jakarta (**β=**1.154; 95% CI, 0.8609 - 1.446). A similar pattern occurred within the AJCUI and Indonesia data (**β=** 1.262; 95% CI, 0.7373 - 1.787), as displayed in **Figure 5b**. Consequently, it is plausible to conclude that the trend of COVID-19 cases in AJCUI is representative of cases reported in DKI Jakarta and Indonesia. This is most likely due to the fact that the AJCUI COVID-19 laboratory is a part of DKI Jakarta and Indonesia. We can expect the positive percent trend will continue to hold true for North Jakarta data and other regions in DKI Jakarta province. In addition, as depicted in **Figure 1**, at the end of 2022, DKI Jakarta data increasingly resemble AJCUI data. From this perspective, an increased positivity rate in the laboratory test could serve as an indicator of a rise in the number of cases in the community.

**Figure 5.**
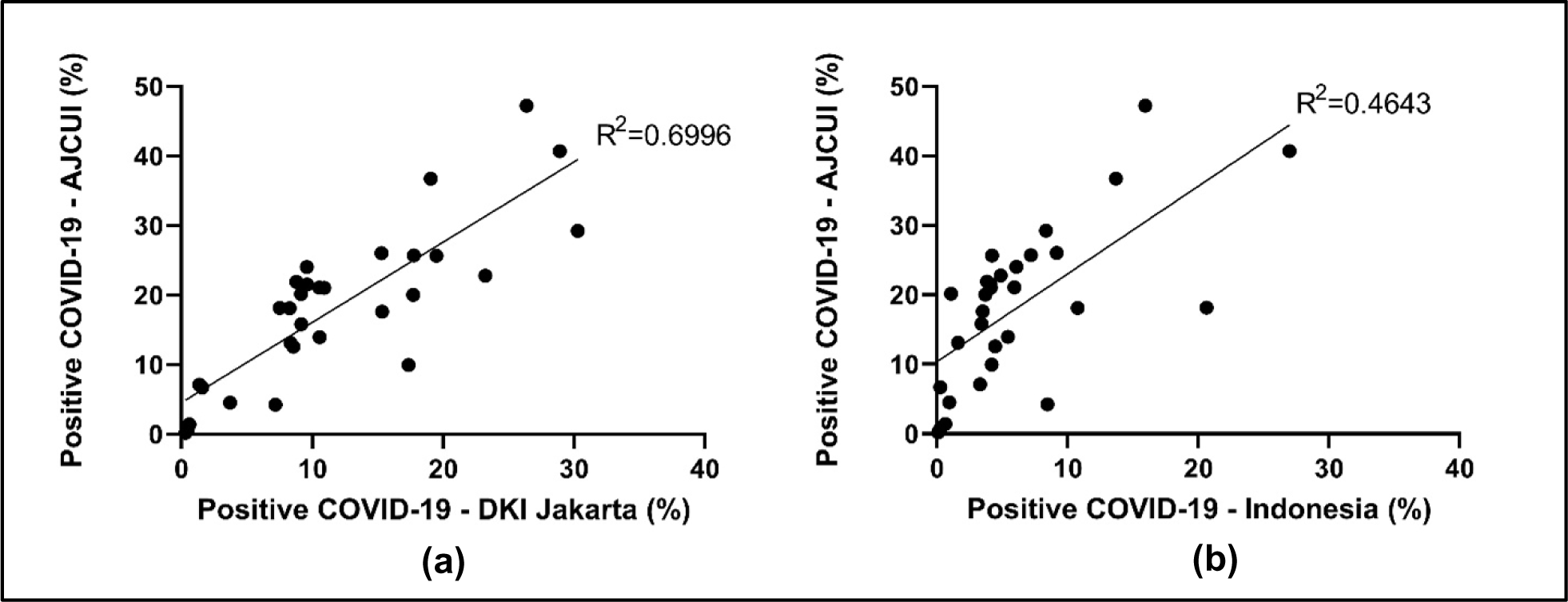
A linear positive correlation of COVID-19 confirmed cases between AJCUI and DKI Jakarta; AJCUI and Indonesia. The dots represent the fitted average positive percentage of COVID-19 from (a) AJCUI and DKI Jakarta data (b) AJCUI and Indonesia data for each represented period from July 2020 to December 2022.

While a positive correlation was observed between AJCUI and DKI Jakarta positivity rates, we further investigated whether the Ct value, which represents viral load, may contribute to the positivity rates for AJCUI and DKI Jakarta. In consideration of the public health emergency caused by the emergence of COVID-19, the Secretary of Health and Human Services authorized a EUA for emergency use of the CDC 2019-Novel Coronavirus Real-Time RT-PCR Diagnostic Panel on February 4, 2020. The FDA issued immediate guidance on February 29, 2020, to facilitate the expansion of available tests and testing facilities. Since then, diagnostic testing has improved globally, including the interpretation of the SARS-CoV-2 test by incorporating the Ct value [42,43]. **Figure 6** displays the correlation between the Ct value in AJCUI and the increase and decrease in the proportion of positive AJCUI cases. When the proportion of Ct value < 30 was more prevalent, COVID-19-positive cases tended to rise. While the proportion of Ct value > 30 is more prevalent, the proportion of positive COVID-19 similarly tends to decline. This occurrence was evident between November 2021 and May 2022. Although the number of positive cases was low, the proportion of Ct values was rising, which was followed by an increase in positive cases within the following month.

**Figure 6.**
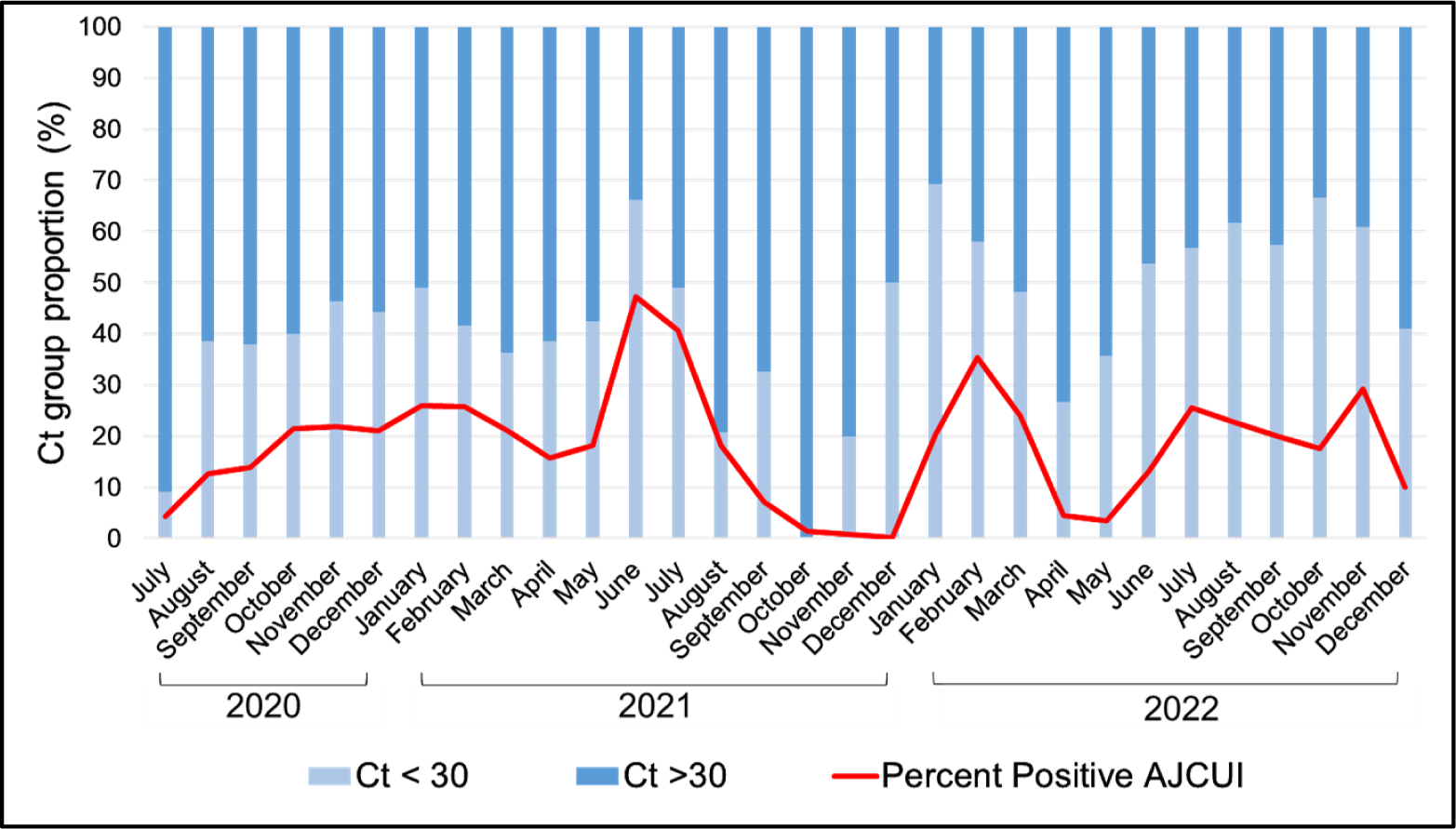
Ct values dynamics regarding positive cases. The graph was based on 30 months of collected data in AJCUI COVID-19 laboratory. Red line represents the fluctuation of the positive percentages of cases monthly. Light blue and dark blue bars show proportion of Cr < 30 and Ct > 30, respectively.

To further analyze the percentage of positive cases with a Ct value greater than 30 and less than 30, we demonstrated the correlation using simple linear regression in **Figure 7**. The Ct value < 30 in AJCUI is positively correlated with the increase in positive percentage (β=0.4416), and vice versa (**Figure 7a** and **Supplementary File 4**). Furthermore, as previously mentioned in **Figure 5**, the AJCUI and DKI Jakarta data, and AJCUI and Indonesia data presented a similar trend and a positive correlation. Consequently, we analyzed the Ct value < 30 proportion of AJCUI concerning the Jakarta positivity rate and Indonesia positivity rate. It demonstrated a similar result of positive correlation of AJCUI and DKI Jakarta (β=0.3188, **Figure 7b**). It is possible that the same trend occurred due to the even distribution of testing coverage in DKI Jakarta. However, AJCUI data may not accurately reflect the positivity rate of Indonesia (β=0.0389, **Figure 7c**) because of the geographical situation, disparity of COVID-19 public information, and uneven testing coverage. The converse correlation was seen when plotting the Ct > 30 from AJCUI data to DKI Jakarta and Indonesia positivity rate (**Supplementary File 4**). From this correlation, AJCUI, DKI Jakarta, and Indonesia drew the same conclusion: that Ct < 30 dominated when the positive rate was high, whereas Ct > 30 dominated when the positivity rate was low. We assume that this trend was limited to Greater DKI Jakarta and not in Indonesia, although a number of studies have demonstrated that Ct values have the potential to serve as an epidemiological early-warning indicator [44–46].

**Figure 7.**
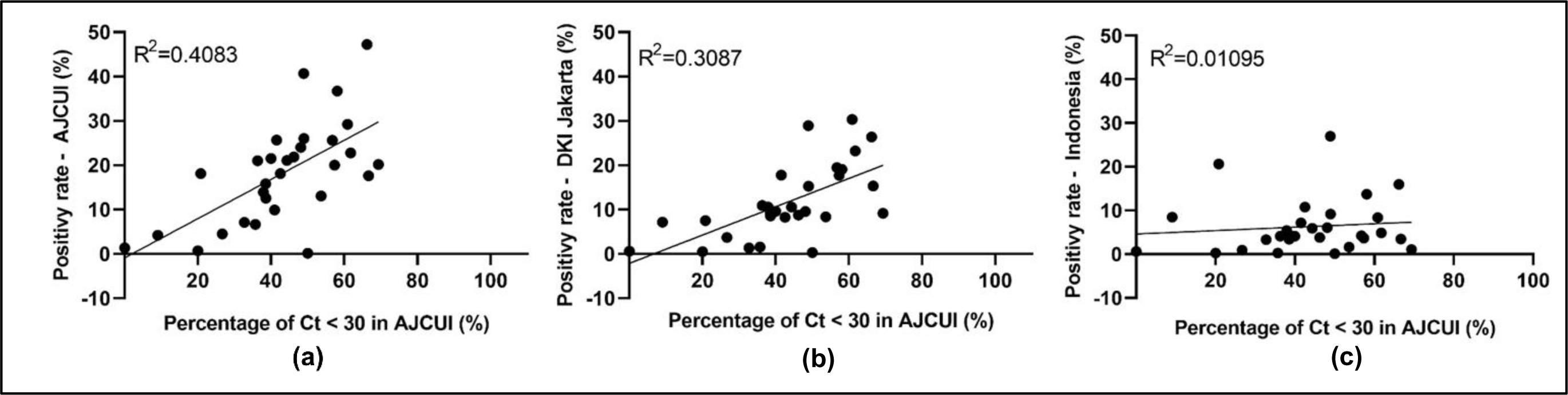
Correlation of positivity rate and high viral intensity (Ct < 30). The dots represent the appropriate positivity rates of (a) AJCUI, (b) DKI Jakarta and (c) Indonesian data to the percentage of COVID-19 confirmed cases data from AJCUI laboratory with the Ct < 30 during the 30-months study period.

During the first and second waves of the pandemic, the population in Madrid experienced a dynamic mean of SARS-CoV-2 RT-PCR Ct values with sustained reduction of viral load (Ct: 23.4-32.3) in the first ten weeks, followed by stability with viral load (Ct: 31.9-35.5) for nine weeks and a sharp increase until reaching stability at high levels in the following twelve weeks; this coincided with an increase in the proportion of positive cases and a decrease in the median age [47]. The relationship between the Ct value and the positive percentage can be explained by conditions in which infected individuals who have a low Ct value also have a higher probability of transmitting the virus, thereby increasing the positivity rate in general, or by the fact that, when the positive percentage is high, the dominant population consists of individuals with a higher viral load and are more infectious. The reverse also applies to a high Ct value with a positive percentage that tends to be low [48].

### 3.6 Vaccination contribution to COVID-19

Indonesia began vaccinations against COVID-19 on January 14, 2021. The second dose was administered mostly a month later, on February 17, 2021. Subsequently, the third dose was started to be distributed on July 17, 2021. The distribution and administration of COVID-19 vaccines in Indonesia vary according to the level of COVID-19 transmission. DKI Jakarta in particular is one of the prioritized provinces due to the high level of transmission. Therefore, we analyze AJCUI’s data restricted to DKI Jakarta as it represents regional data of this province [49,50]. The vaccination rate for the first two doses increased instantly because they were mandatory before people could enter public spaces. This policy was enforced by scanning QR codes with the PeduliLindungi application. The third dose or first booster vaccination coverage was slower. However, the third dose was gradually increased after it was mandated, particularly for travel, starting at the beginning of 2022. Vaccine distribution was prioritized for front-line workers, the elderly, adults and young adults, kids and teenagers (6–11 years old), nursing mothers, and toddlers older than 2 years old, subsequently. The most widely distributed vaccines in Indonesia are the Sinovac/CoronaVac vaccines, which are mostly used for the first and second doses. Both of the vaccines were considered to be less effective (50% efficacy) than those manufactured by Pfizer (95% efficacy), Moderna (94% efficacy), and AstraZeneca (70% efficacy) [51].

In the case of COVID-19, vaccines are not intended to prevent infection; rather, they alleviate the severity of the infection. Therefore, the mortality could be lessened due to being clinically less severe for the infected patients. Furthermore, even though SARS-CoV-2 keeps on mutating to evade the immune system or even herd immunity, asymptomatic cases become more common for people who are already vaccinated, either by being previously infected or receiving vaccines. Therefore, survivability is expected to increase. Even more, several studies indicate that vaccines can make infected individuals less contagious [52,53].

**Figure 8** demonstrates that vaccination coverage does not alleviate the emergence of the case positivity peak whenever a new virus variant emerges. Viruses that mutate into new variants and sub-variants can infect both vaccinated and unvaccinated COVID-19-infected individuals. Despite the coverage of the first, second, and third vaccine doses, the AJCUI and DKI Jakarta positivity rate patterns are similarly fluctuating in response to the emergence of new variants. **Figure 9** depicts that the Delta variant resulted in the highest mortality rate during the second wave to strike DKI Jakarta. Although the third vaccine was being distributed, uptake was extremely slow, and hence the lesser mortality peak associated with the Omicron variety occurred once again. This was due to the virus continuing to mutate, and it was possible that the vaccinated population had not yet been affected or had a lower mortality rate, as reported by numerous studies [54–56]. Overall, the duration of vaccination administration is correlated with the number of deaths in consideration of the virus variant.

**Figure 8.**
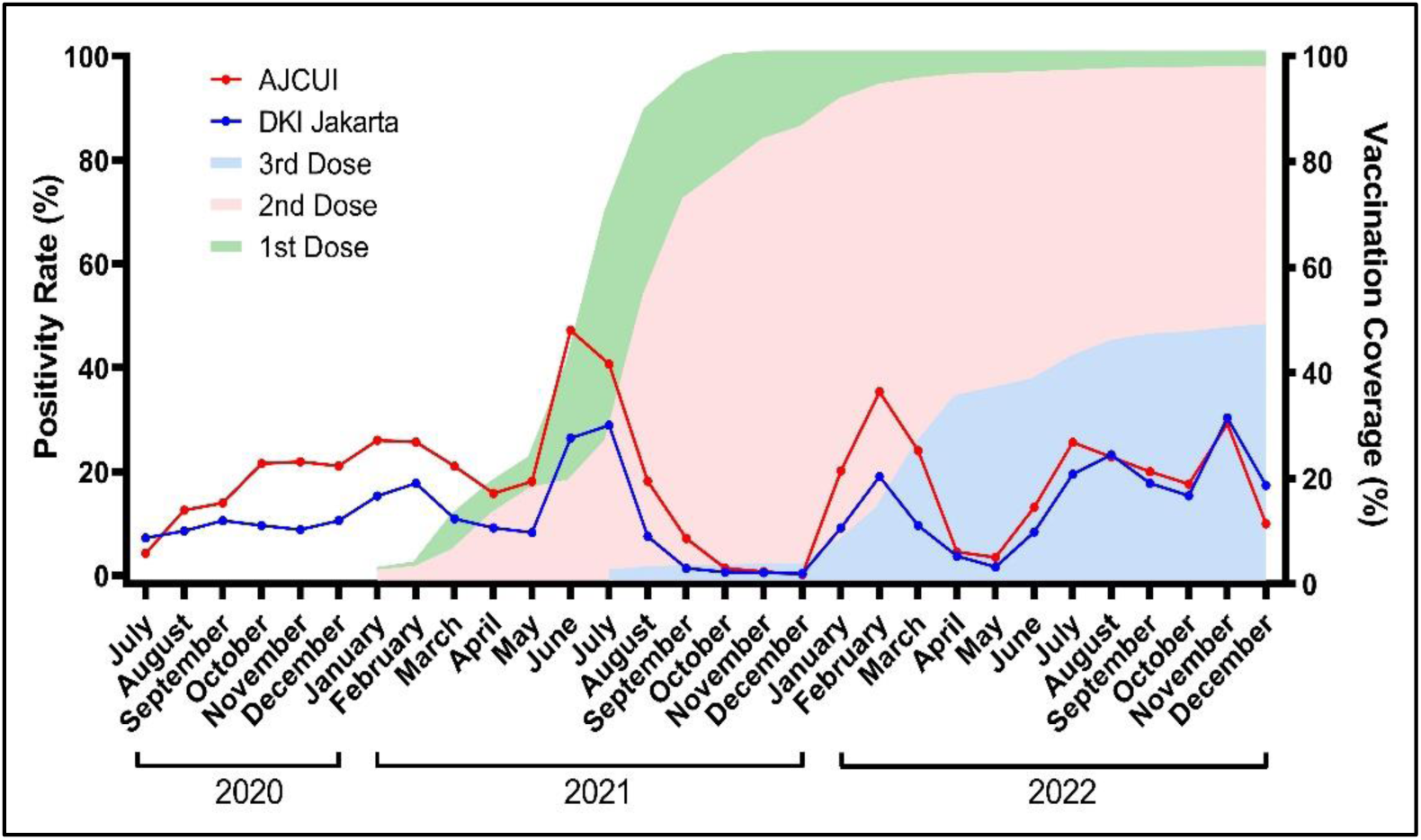
Vaccination and positivity rate. Coverage of the first (green shaded area), second (pink shaded area) and third (light blue shaded area) of DKI Jakarta COVID-19 vaccination (https://vaksin.kemkes.go.id/#/vaccines) in correlation with positivity rate of AJCUI (red line) and DKI Jakarta (blue line) during the 30-months study period.

**Figure 9.**
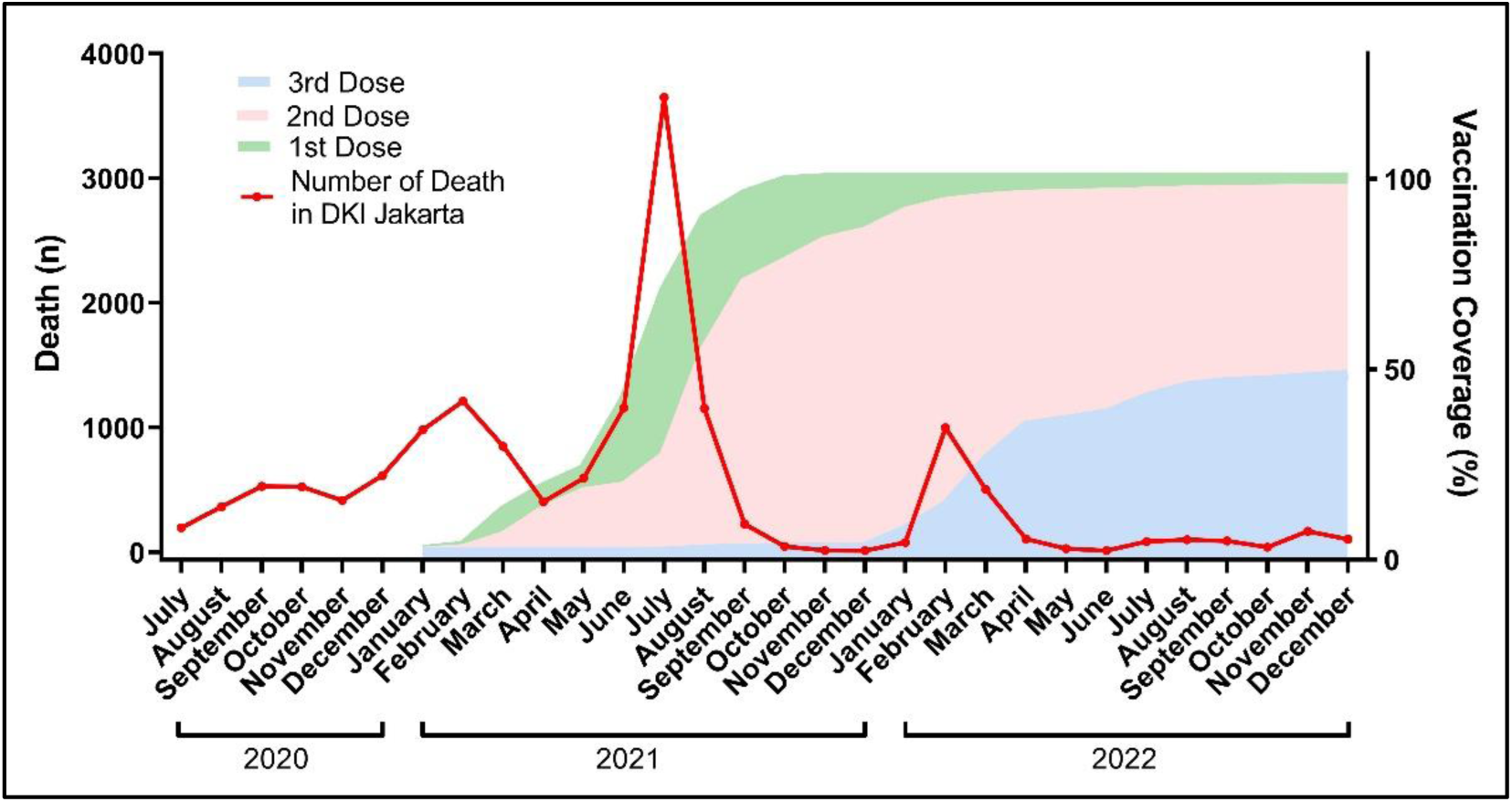
Vaccination and death from COVID-19. First (green shaded area), second (pink shaded area) and third (light blue shaded area) of COVID-19 vaccination (https://vaksin.kemkes.go.id/#/vaccines) percentage in correlation with the number of deaths (red line) in DKI Jakarta (https://corona.jakarta.go.id/id) during the 30-months study period.

## 4. Conclusions

During the 30 months of the global pandemic, which impacted various sectors, including the improvement of diagnostic detection, the AJCUI COVID-19 reference laboratory presented comprehensive data demonstrating the similar trend of COVID-19 prevalence with DKI Jakarta and Indonesia, as well as contrasting testing and positivity rates relative to the parallel data from these two designated areas. AJCUI COVID-19 laboratory data indicate that age groups with high mobility, ages 20 to 50, dominated testing and positive cases. The policies applied in DKI Jakarta and Indonesia are consistent with WHO guidelines and capable of containing the epidemic for two years. Government policy has an essential impact on reducing the number of positive cases in the country. In addition, our study interprets the SARS-CoV-2 test by integrating the Ct value in order to better comprehend the frequency of the Ct value in the positive case group. Data from AJCUI and DKI Jakarta, as well as data from AJCUI and Indonesia, exhibited a similar trend and a positive correlation. Further study of Ct values has the potential to serve as an early warning signal for an anticipated increase in positive cases. In response to the emergence of new variants, the AJCUI and DKI Jakarta positive rate patterns fluctuate identically despite the coverage of the first, second, and third vaccination doses. Generally, the period of vaccination administration is correlated with the mortality rate when the viral variant is considered.

## Data Availability

All data produced in the present study are available upon reasonable request to the authors

## Credit authorship contribution statement

**MMMK:** Conceptualization, Methodology, Data collection, Data management, Data verification, Data analysis, Supervision, Data Visualization, Writing the manuscript, Review.

**TAW, SJ, HK:** Methodology, Data collection, Data management, Data extraction, Data analysis, Data Visualization, Writing original draft.

**SSS, V, JWW:** Data collection and Data management, Review.

**ET, ADVT:** Data collection, Data verification, Review.

**IM:** Data analysis, Review.

**SA:** Conceptualization, Methodology, Supervision, Data analysis, Writing the manuscript, Review.

## Acknowledgement

We would like to thank the distinguished head of faculty members for their support during the COVID-19 Laboratory AJCUI operation: Felicia Kurniawan. MD, Prof. Yuda Turana., MD, Stefanus Lembar., MD (alm), Sheella R. Bororing., MD, Lilis., MD, and Mrs. Debora Yuliawati. We appreciate the help from our former COVID-19 laboratory staff Isadora G. Sahusilawane dentistry, Linawati Hannanta., MD, and Meiliyana Wijaya., MD. Also, to our collaborating sites for specimen collections: School of Medicine and Health Sciences, AJCUI, and Atma Jaya Teaching Hospital. We would also like to acknowledge all the participating healthcare personnel and volunteers for contributing to specimen processing and our COVID-19 laboratory technicians: Sri Handayani, Muhammad Yashir, Ariska V. Marantika, Ida Afiyah, Fitriani Nurhasanah, and Linda Widyawati.

---

**Supplementary File 1.** Government policies during the COVID-19 pandemic in Indonesia.

| Policies | Date of releases | Reference | Date of Implementation | Detail of the policies |
| --- | --- | --- | --- | --- |
| Large-Scale Social Restrictions (LSSR) | 9 April 2020 | Governor Regulation of DKI Jakarta Number 33 of 2020 regarding LSSR Implementation | 5 June - 13 September 2020 | Phase 1<br>• Schools are conducted online<br>• Office capacity 50%<br>• Worship place capacity 50%<br>• Mall and shopping center capacity 50%<br>• Personal cars capacity 50%<br>• Public transport capacity 50%<br>• Even Odd policy is enforced<br>• COVID-19-positive - self-isolation is allowed to be done at home |
|  |  |  | 14 September - 11 October 2020 | Transition Phase 2<br>• Visitor registration for contact tracing in public places<br>• Even Odd policy is not applicable<br>• Swimming pool, gym, cinema capacity 25% complying to health protocols<br>• Dine-ins are allowed in restaurants with 1.5-2 meters of seating distance unless living under one roof |
|  |  |  | 12 October - 10 January 2021 | Phase 2<br>• School is conducted online<br>• Office capacity 25% |
|  |  |  |  | • Worship place capacity 50% (those located in red zone must be closed)<br>• Mall and shopping center capacity 50%<br>• Personal cars capacity max. 2 person /row<br>• Public transport capacity 50%<br>• Even Odd policy is not applicable<br>• Self isolation must be done in wisma atlet / referenced hotel and lodging<br>• No dine-in allowed in restaurants |
|  |  |  | 11 January - 8 February 2021 | Restricted<br>• Office capacity 25% WFO<br>• School is conducted online<br>• Essential and construction sector capacity 100% complying with health<br>• The shopping center closes at 19.00<br>• Dine-ins are allowed in restaurants max. 18.00 with 25% capacity; takeaways are allowed 24 hours as normal operational hours<br>• Worship place capacity 50%<br>• Cultural sites and public places are closed<br>• Medical facility capacity 100% complying to health protocols |
|  |  |  |  | Public transport capacity 50% |
| Micro-scale Activity Restrictions | 8 February 2021 | Instruction of Home Affairs Minister Number 03 of 2021 | 9 February - 2 July 2021 | Office located in red zones capacity 25% & 50% for those in other zones<br>Schools located in red zones conducted online & those located in other zones are following Kemendikbud Ristek policies<br>Essential sector capacity 100% Dine-ins capacity 25% & takeaways are allowed 24 hours as normal operational hours<br>Shopping center capacity 25% & closes at 20.00<br>Construction site capacity 100%<br>Worship place located in the red zone following SE Menag & other zones following Kemenag policy<br>Activities in public space in red zones are canceled & other zones are allowed at 25%<br>Public transportation capacity 50% |
| Emergency Community Activities Restrictions Enforcement (CARE) | 1 July 2021 | Instruction of Home Affairs Minister Number 15 of 2021 | 3 - 25 July 2021 | Non-essential sector WFH 100%, essential sector WFH 50%, critical sector WFO 100%<br>Shopping centers are closed<br>Take-away and delivery only for restaurants<br>Public transportation capacity 70%<br>Worship places are closed |
|  |  |  |  | Public space/activities are closed/canceled<br>The shopping center for essential needs closes at 20.00 with a 50% capacity |
| Level 4<br>CARE | 20 July 2021 | Instruction of Home Affairs Minister Number 24 of 2021 | 26 July - 25 August 2021 | School is conducted online<br>Non-essential sector 100% WFH<br>Essential sector 25% WFO<br>Critical sector 100% WFO<br>Restaurant with open area capacity 25% capacity max. 30 minutes /table<br>Shopping center capacity 50% + Peduli Lindungi registrations<br>Entertainment (cinema, karaoke) is closed<br>Construction activities 100%<br>Public facilities are closed (parks, public areas)<br>Public transport capacity 50%<br>No wedding receptions are allowed |
| Level 3<br>CARE | 25 August 2021 | Instruction of Home Affairs Minister Number 26 of 2021 | 26 August - 30 August 2021 | In-person schools are allowed for 50% capacity<br>Shopping center capacity 50% + Peduli Lindungi registrations<br>Restaurants to serve dine-in for max. 30 minutes/table<br>Public transportation capacity 70%<br>Weddings are allowed for 20 invitees without dine-ins |
| Level 2<br>CARE | 25<br>August<br>2021 | Instruction of Home<br>Affairs Minister<br>Number 26 of 2021 | 31 August - 30 December<br>2022 (Varies according to<br>the number of the positive<br>case) | • School is conducted 50:50 (PTM Terbatas)<br>• All sectors 75% WFO<br>• All kinds of public places at 75% capacity, closed at 20.00+<br>Peduli Lindungi registrations<br>• Weddings and cultural ceremonies are allowed 75%<br>capacity with proper health protocols |
| Level 1<br>CARE | 25<br>August<br>2021 | Instruction of Home<br>Affairs Minister<br>Number 26 of 2021 | 31 August - 30 December<br>2022 (Varies according to<br>the number of the positive<br>case) | • School is conducted 50:50 (PTM Terbatas)<br>• All sectors 75% WFO<br>• All kinds of public places at 75% capacity, closed at 22.00<br>+ Peduli Lindungi registrations<br>• Restaurants opening later in the day, allowed to open until<br>00.00<br>• Weddings and cultural ceremonies are allowed 75%<br>capacity with proper health protocols |

**Supplementary File 2.** The number of samples, positive cases and positivity extracted from AJCUI, DKI Jakarta and Indonesia.

| Year | Month | AJCUI |  |  | DKI Jakarta |  |  | Indonesia |  |  |
| --- | --- | --- | --- | --- | --- | --- | --- | --- | --- | --- |
|  |  | Number of |  | Positivity Rate (%) | Number of |  | Positivity Rate (%) | Number of |  | Positivity Rate (%) |
|  |  | Sample | Positive |  | Sample | Positive |  | Sample | Positive |  |
| 2020 | July | 1085 | 46 | 4.24 | 231971 | 16676 | 7.19 | 1281005 | 108376 | 8,46 |
|  | August | 3318 | 417 | 12.57 | 229936 | 19697 | 8.57 | 1492000 | 66420 | 4,45 |
|  | September | 7620 | 1061 | 13.92 | 328778 | 34743 | 10.57 | 2065020 | 112212 | 5,43 |
|  | October | 5290 | 1138 | 21.51 | 316670 | 30414 | 9.60 | 2998014 | 123080 | 4,11 |
|  | November | 3909 | 856 | 21.90 | 359679 | 31606 | 8.79 | 3350000 | 128795 | 3,84 |
|  | December | 6732 | 1418 | 21.06 | 471356 | 49734 | 10.5 | 3445025 | 204315 | 5,93 |
| 2021 | January | 7861 | 2047 | 26.04 | 558316 | 85383 | 15.29 | 3656060 | 335116 | 9,17 |
|  | February | 4144 | 1064 | 25.68 | 385403 | 68461 | 17.76 | 3571018 | 256320 | 7,18 |
|  | March | 4517 | 950 | 21.03 | 381073 | 41599 | 10.92 | 4303022 | 177078 | 4,12 |
|  | April | 2481 | 392 | 15.80 | 285225 | 26154 | 9.17 | 4601011 | 156656 | 3,40 |
|  | May | 1260 | 228 | 18.10 | 254251 | 21032 | 8.27 | 1424075 | 153335 | 10.77 |
|  | June | 5114 | 2415 | 47.22 | 456215 | 120436 | 26.40 | 2235324 | 356569 | 15.95 |
|  | July | 6467 | 2630 | 40.67 | 920865 | 266347 | 28.92 | 4562966 | 1231386 | 26.99 |
|  | August | 1588 | 288 | 18.14 | 450977 | 33903 | 7.52 | 3294754 | 680143 | 20.64 |
|  | September | 773 | 55 | 7.12 | 487958 | 6660 | 1.36 | 3801250 | 125303 | 3.30 |
|  | October | 722 | 10 | 1.39 | 614991 | 3707 | 0.60 | 4600532 | 29254 | 0.64 |
|  | November | 703 | 5 | 0.71 | 472562 | 2395 | 0.51 | 4803168 | 12051 | 0.25 |
|  | December | 1192 | 2 | 0.17 | 420256 | 1398 | 0.33 | 5800984 | 6311 | 0.11 |
| 2022 | January | 1295 | 261 | 20.15 | 593422 | 54328 | 9.16 | 8392751 | 90650 | 1.08 |
|  | February | 3228 | 1185 | 36.71 | 1357914 | 258730 | 19.05 | 8831218 | 1211078 | 13.71 |
|  | March | 1004 | 241 | 24.00 | 624608 | 59904 | 9.59 | 7380228 | 448370 | 6.08 |
|  | April | 662 | 30 | 4.53 | 253597 | 9438 | 3.72 | 3500365 | 33145 | 0.95 |
|  | May | 403 | 27 | 6.70 | 192044 | 3065 | 1.60 | 3022454 | 7759 | 0.26 |
|  | June | 626 | 82 | 13.10 | 238923 | 19961 | 8.35 | 2082087 | 33487 | 1.61 |
|  | July | 749 | 192 | 25.63 | 443322 | 86382 | 19.49 | 2817542 | 118638 | 4.21 |
|  | August | 803 | 183 | 22.79 | 465331 | 108074 | 23.23 | 2908378 | 141594 | 4.87 |
|  | September | 610 | 122 | 20.00 | 269636 | 47769 | 17.72 | 1737990 | 64192 | 3.69 |
|  | October | 545 | 96 | 17.61 | 210399 | 32268 | 15.34 | 1583333 | 55233 | 3.49 |
|  | November | 1006 | 294 | 29.22 | 315320 | 95569 | 30.31 | 1759480 | 146926 | 8.35 |
|  | December | 1055 | 105 | 9.95 | 177607 | 30824 | 17.36 | 1347797 | 56520 | 4.19 |

**Supplementary File 3.** The number of samples tested and the positive cases percentage in each age group from AJCUI data.

| Year | Month | Age Group<br>Total Number (% of Positive Cases) |  |  |  |  |  |  |
| --- | --- | --- | --- | --- | --- | --- | --- | --- |
|  |  | 0-10 | 11-20 | 21-30 | 31-40 | 41-50 | 51-60 | >60 |
| 2020 | July | 7 (28.6) | 49 (0) | 376 (2.7) | 251 (3.2) | 222 (5.9) | 117 (8.5) | 59 (6.8) |
|  | August | 33 (12.1) | 134 (8.2) | 844 (10.3) | 876 (10.8) | 705 (13.2) | 408 (16.4) | 255 (25.1) |
|  | September | 113 (15.9) | 227 (15.4) | 2086 (12.1) | 2045 (14.1) | 1486 (13.1) | 866 (19.2) | 544 (19.5) |
|  | October | 65 (24.6) | 169 (17.2) | 1600 (21) | 1373 (20.4) | 1017 (23.2) | 631 (22) | 395 (25.8) |
|  | November | 79 (25.3) | 169 (26.6) | 1137 (17.3) | 1004 (20.4) | 671 (21) | 485 (29.5) | 345 (30.1) |
|  | December | 146 (23.3) | 366 (19.7) | 2015 (19.5) | 1625 (20.4) | 1114 (21.5) | 758 (24.7) | 615 (24.9) |
| 2021 | January | 183 (26.8) | 416 (25.2) | 2389 (23.3) | 1871 (27) | 1302 (27) | 833 (29.4) | 796 (29.4) |
|  | February | 86 (37.2) | 192 (29.2) | 1365 (19.9) | 952 (28.4) | 651 (24.4) | 430 (29.3) | 455 (33) |
|  | March | 96 (34.4) | 185 (30.3) | 1213 (17.8) | 1156 (19.4) | 834 (19.1) | 432 (29.9) | 406 (32.8) |
|  | April | 53 (32.1) | 74 (18.9) | 758 (12.1) | 665 (13.7) | 521 (15) | 234 (23.1) | 172 (26.7) |
|  | May | 24 (33.3) | 94 (12.8) | 399 (13.3) | 260 (23.8) | 205 (17.6) | 132 (15.2) | 143 (25.2) |
|  | June | 132 (49.2) | 294 (41.5) | 1533 (47.3) | 1196 (48.2) | 856 (49.3) | 580 (50) | 510 (42.4) |
|  | July | 189 (52.9) | 406 (42.4) | 1719 (36.8) | 1379 (38) | 1116 (44.4) | 778 (42.9) | 863 (42.6) |
|  | August | 28 (32.1) | 88 (30.7) | 499 (14.4) | 262 (14.9) | 287 (16.4) | 217 (20.7) | 203 (24.1) |
|  | September | 7 (14.3) | 48 (18.8) | 273 (5.1) | 122 (8.2) | 110 (2.7) | 89 (1.1) | 102 (17.6) |
|  | October | 8 (0) | 58 (0) | 293 (1.4) | 103 (1) | 95 (0) | 64 (0) | 75 (6.7) |
|  | November | 11 (9.1) | 43 (0) | 269 (0.4) | 78 (1.3) | 94 (0) | 61 (1.6) | 120 (0.8) |
|  | December | 192 (0.5) | 412 (0) | 197 (0) | 90 (1.1) | 69 (0) | 72 (0) | 98 (0) |
| 2022 | January | 104 (6.7) | 95 (28.4) | 480 (15.2) | 175 (33.7) | 116 (25) | 108 (25.9) | 148 (25.7) |
|  | February | 154 (31.2) | 261 (37.2) | 1000 (36.3) | 605 (37.7) | 468 (35) | 313 (35.5) | 415 (40.7) |
|  | March | 93 (10.8) | 86 (25.6) | 309 (24.6) | 119 (27.7) | 115 (29.6) | 92 (20.7) | 177 (24.9) |
|  | April | 89 (6.7) | 52 (1.9) | 130 (5.4) | 84 (8.3) | 51 (11.8) | 52 (1.9) | 108 (1.9) |
|  | May | 7 (0) | 39 (5.1) | 105 (17.1) | 57 (0) | 43 (7) | 57 (0) | 94 (4.3) |
|  | June | 9 (33.3) | 25 (8) | 220 (9.1) | 109 (26.6) | 69 (20.3) | 58 (10.3) | 128 (6.3) |
|  | July | 33 (54.5) | 51 (25.5) | 226 (27) | 131 (26) | 95 (24.2) | 79 (24.1) | 134 (17.9) |
|  | August | 40 (57.5) | 52 (34.6) | 232 (19) | 132 (25.8) | 109 (15.6) | 85 (20) | 153 (19.6) |
|  | September | 28 (39.3) | 51 (25.5) | 165 (19.4) | 91 (23.1) | 73 (17.8) | 79 (16.5) | 120 (14.2) |
|  | October | 22 (27.3) | 24 (20.8) | 180 (12.2) | 81 (29.6) | 62 (16.1) | 66 (15.2) | 107 (17.8) |
|  | November | 50 (38) | 138 (18.1) | 326 (24.2) | 148 (40.5) | 85 (31.8) | 79 (31.6) | 180 (47.8) |
|  | December | 24 (20.8) | 591 (2.4) | 127 (16.5) | 77 (29.9) | 61 (13.1) | 52 (17.3) | 123 (20.3) |

**Supplementary File 4.**
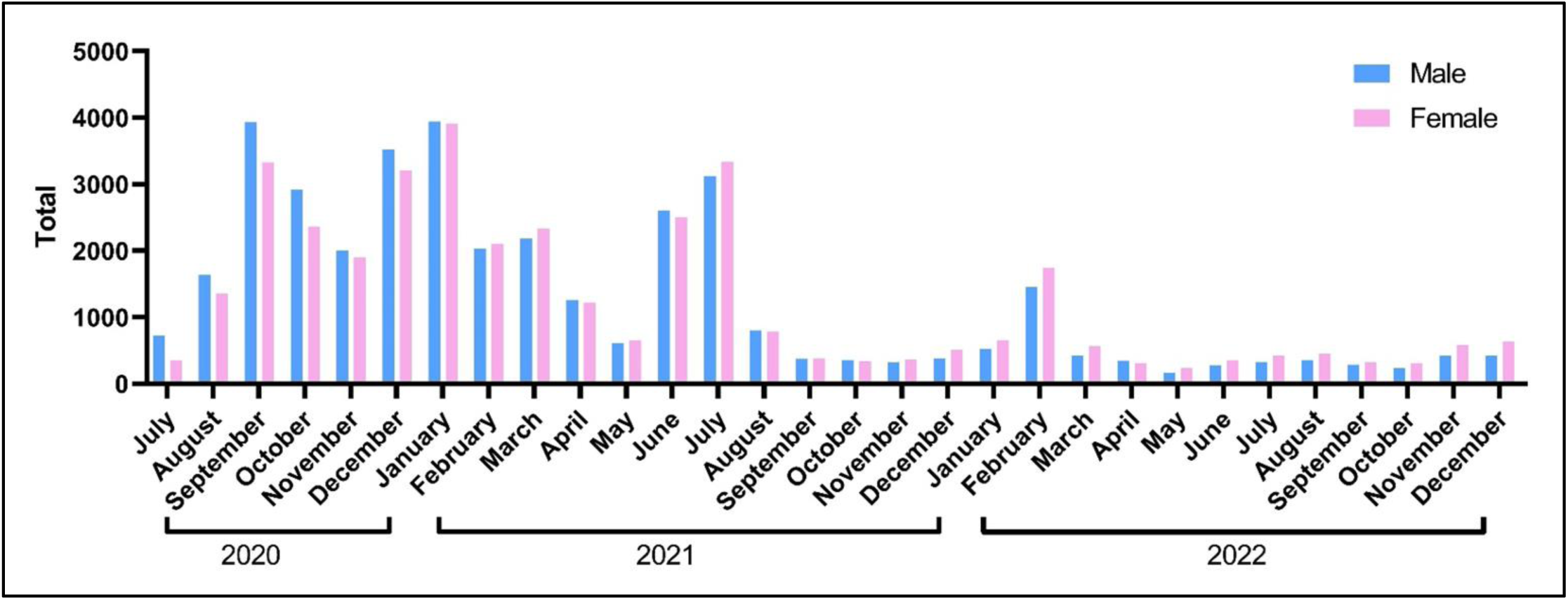
The proportion of total number of gender stratification. The data from the AJCUI COVID-19 laboratory on samples provided for period of July 2020 to December 2022.

**Supplementary File 5.**
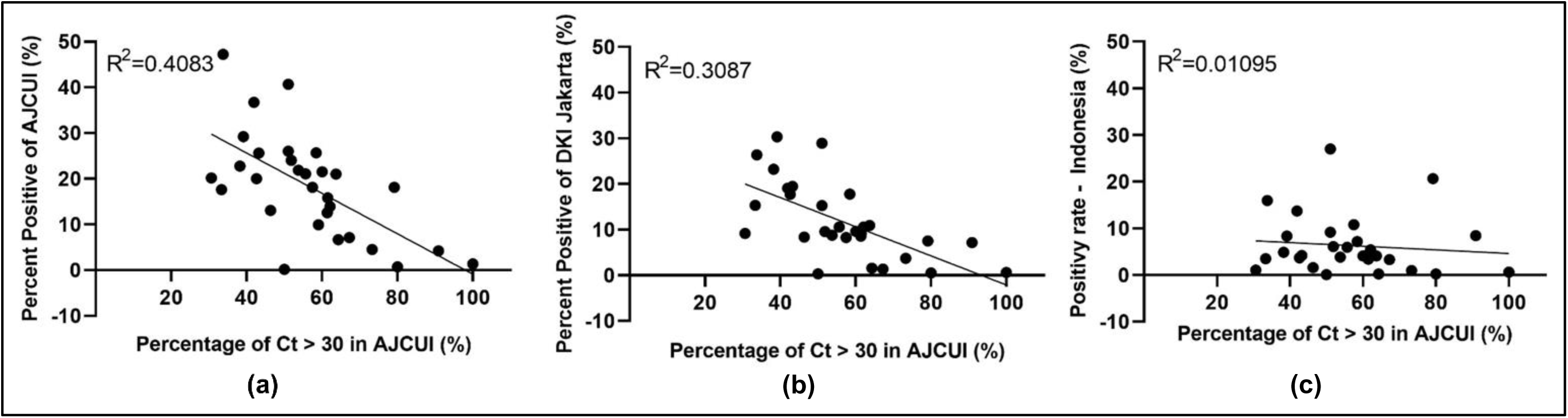
Correlation of positivity rate and low viral intensity (Ct > 30). The dots represent the appropriate positivity rates of (a) AJCUI, (b) DKI Jakarta and (c) Indonesian data to the percentage of COVID-19 confirmed cases data from AJCUI laboratory with the Ct > 30 during the 30-months study period.

